# Olanzapine, Risperidone and Clozapine prescribing is associated with increased risk for Alzheimer’s Disease reflecting antipsychotic-specific effects on microglial phagocytosis

**DOI:** 10.1101/2023.11.10.23298358

**Authors:** Mrityunjoy Mondal, Shiden Solomon, Jiangwei Sun, Nirmal Kumar Sampathkumar, Ivo Carre, Marie-Caroline Cotel, Puja R. Mehta, Lawrence Rajendran, Anthony C. Vernon, Fang Fang, Jacqueline Mitchell

**Author notes:** Deutsches Zentrum für Neurodegenerative Erkrankungen e. v. (DZNE), Bonn, Germany. Alzheimer’s Research UK Oxford Drug Discovery Institute, Centre for Medicines Discovery, University of Oxford, Oxford, United Kingdom.

## Abstract

Epidemiological data provides evidence for a positive correlation between schizophrenia diagnosis and an increased risk to develop dementia. Whether and how use of antipsychotic medication may contribute to this association is however unknown. We therefore conducted a pharmaco-epidemiological study based on Swedish Patient and Prescribed Drug Registers to investigate the effect of three antipsychotics, Olanzapine, Risperidone, and Clozapine, on dementia risk. Our data suggest that prescription of all three antipsychotics is significantly associated with increased risk of Alzheimer’s disease (AD) and other dementias including vascular dementia. To provide a nexus of causality to this association, we explored the impact of these drugs on microglia and neurons using cells derived from human induced pluripotent stem cells (hiPSCs). Acute exposure to Olanzapine and Risperidone did not significantly alter amyloid-β (Aβ) production in hiPSC-derived cortical neurons, but suppressed hiPSC-derived microglial-mediated Aβ clearance, leading to Aβ accumulation. Neither Olanzapine nor Risperidone had any significant effect on hiPSC-derived microglial synaptosome phagocytosis. Conversely, Clozapine significantly reduced Aβ production in neurons, and increased microglial uptake of Aβ but also synaptosomes, consistent with higher lysosomal levels in Clozapine-exposed hiPSC-derived microglia. These data provide the first evidence that antipsychotics prescribed to individuals with schizophrenia are associated with increased risk for dementia and suggest potential cellular bases for this effect via the modulation of microglia uptake of Aβ and synapses in a drug specific manner.

## Introduction

Dementia prevelance is rapidly increasing in the aging global population (<u>Weller and Budson, 2018</u>), with Alzheimer’s disease (AD) and vascular dementia constituting the majority of cases (60% and 20% respectively) (<u>Rizzi et al., 2014</u>). Aside from declines in cognitive performance and memory, dementia is also commonly associated with other behavioural and psychological symptoms, complicating its management (<u>Cerejeira et al., 2012</u>). It is estimated that up to 90% of AD patients will develop at least one symptom of psychosis in the course of their illness (<u>Liperoti et al., 2008</u>).

Antipsychotics are primarily prescribed to treat psychosis including schizophrenia, bipolar disorder and other psychotic disorders, as well as psychiatric and behavioural changes in dementia (<u>Tifratene et al., 2017</u>). However, antipsychotic drugs including the dopamine D2 and serotonin 5HT2 receptor antagonists Olanzapine, Risperidone, and Clozapine are associated with numerous side effects including sedation and weight gain and their efficacy to improve negative symptoms and cognition is debated (<u>Dayabandara et al., 2017</u>). Of note, anticholinergic antipsychotics are also linked with an increased risk of developing dementia (<u>Coupland et al., 2019</u>).

Antipsychotic induced side effects increase with age, and long-term use of these treatments may exacerbate dementia-related symptoms (<u>Hulshof et al., 2015</u>). A systematic review found that the use of atypical antipsychotics to treat non-cognitive symptoms in AD has limited efficacy at best, with negative impacts outweighing the benefits (<u>Schneider et al., 2006</u>). To date, the cellular processes by which antipsychotics may impact AD progression are under-studied. Understanding how antipsychotics impact on dementia disease risk and progression is crucial, not only for the development of strategies to mitigate disease risk but also to improve our knowledge of the cellular processes underlying neurodegenerative disease. In this study, we demonstrate a clear increased risk of AD associated with the use of antipsychotics, and suggest that these drugs may impact the dementia process via alterations in amyloid clearance and synapse engulfment by microglia.

## Results

### Pharmacoepidemiological data suggest an elevated risk of dementia associated with antipsychotic prescribing

To assess the risk for diagnosis of dementia in relation to the use of three different antipsychotics, we performed a nationwide nested case-control study using the Swedish Patient and Prescribed Drug Registers. This included 85,721 patients with newly diagnosed dementia between July 1, 2008 and December 31, 2013, and up to five controls per case that were randomly selected from the general Swedish population and individually matched to the index case by sex, birth year, and area of residence. Information on antipsychotic use was extracted from the Prescribed Drug Register for both cases and controls. A conditional logistic regression model was applied to calculate odds ratios (ORs) and 95% confidence intervals (CIs). As the Swedish Patient Register includes clinical diagnoses through specialist care alone, there is likely a delay between the first clinical diagnosis of dementia and the first specialist care visit, hence we compared antipsychotic use before the register-based diagnosis of dementia and found individuals with dementia had more often taken antipsychotics during all years before the index date (date of diagnosis for cases and date of selection for controls), compared with controls (STable 1). To reduce the concern of delayed registration of dementia in the Patient Register, we used a lag time of 3 years, taking into account antipsychotic prescriptions made more than 3 years prior to the index date in the main analysis. In the main analysis, use of Olanzapine, Risperidone, and Clozapine were all significantly associated with a higher risk of developing dementia (OR=2.38; 95% CI=2.14-2.64 for Olanzapine, OR=2.80; 95% CI=2.60-3.02 for Risperidone, and OR=5.03, 95% CI=3.66-6.92 for Clozapine) (Table 1). Results from two sensitivity analyses using a lag time of 4 or 5 years are similar (STable 2). Positive associations were also observed when breaking down the use of antipsychotics into 3 groups by tertile distributions of the cumulative defined daily doses (Table 1). These associations were stronger among male and younger individuals (<70 years), compared with female and older individuals (≥70 years) (STable 3). In terms of subtypes of dementia, the associations were stronger for vascular dementia and other dementias, compared with AD (STable 4). Because schizophrenia and bipolar disorders are the main indicating conditions for the studied antipsychotics, we performed a stratified analysis to separately analyze individuals with or without a diagnosis of schizophrenia or bipolar disorder. The positive associations between the studied antipsychotics and a higher risk of dementia were noted among individuals both with and without a diagnosis of either schizophrenia or bipolar disorders (STable 5). We also explored the diagnoses of other psychotic disorders among individuals using the studied antipsychotics but had no diagnosis of schizophrenia or bipolar disorders. We found that from the 3 years before the first dispense to 3 years after the last dispense of antipsychotics, these individuals had received diagnoses of anxiety disorders, depressive disorders, substance use disorders, etc. (STable 6).

**Table 1.** Association between use of antipsychotics and risk of any dementia, a nationwide nested case-control study in Sweden, July 2008-December 2013*.

| Use of antipsychotics | Olanzapine |  | Risperidone |  | Clozapine |  |
| --- | --- | --- | --- | --- | --- | --- |
|  | No. of<br>Cases/Controls | OR (95% CI) | No. of<br>Cases/Controls | OR (95% CI) | No. of<br>Cases/Controls | OR (95% CI) |
| No use | 85209/425525 | 1.00(reference) | 84605/424597 | 1.00(reference) | 85645/426523 | 1.00(reference) |
| Use | 512/1074 | 2.38(2.14-2.64) | 1116/2002 | 2.80(2.60-3.02) | 76/76 | 5.03(3.66-6.92) |
| Low cDDD | 167/380 | 2.19(1.83-2.63) | 469/832 | 2.84(2.53-3.18) | 29/22 | 6.55(3.76-11.40) |
| Medium cDDD | 175/347 | 2.51(2.09-3.01) | 294/502 | 2.94(2.54-3.39) | 26/25 | 5.24(3.03-9.08) |
| High cDDD | 170/347 | 2.45(2.04-2.94) | 353/668 | 2.66(2.33-3.02) | 21/29 | 3.66(2.09-6.42) |
| P for trend |  | <0.0001 |  | <0.0001 |  | <0.0001 |

Abbreviations: cDDD, cumulative defined daily dose; CI, confidence interval; OR, odds ratio.
\*Analysis included only antipsychotics dispensed more than 3 years before the index date (date of diagnosis for cases and date of selection for controls). Analyses were adjusted for age, sex and area of residence (matching factors).

### Clozapine treatment alters neuronal Aβ secretion

Our pharmaco-epidemiological data suggest that prescription of Risperidone, Olanzapine and Clozapine is associated with an increased risk of developing dementia, irrespective of the indication for which these drugs were prescribed and the dose or duration of use. To identify a possible cause for this association, we next investigated potential cellular and molecular mechanisms that may drive this increased risk, focusing on AD linked pathological mechanisms.

We acutely (48h) exposed hiPSC-derived cortical neurons to Risperidone, Olanzapine or Clozapine, at a concentration matching the therapeutic plasma levels of these drugs (Figure 1A). Neither Risperidone nor Olanzapine altered Aβ40 secretion (Figure 1B). In contrast, Clozapine significantly reduced extracellular Aβ levels, relative to vehicle controls (Figure 1A). These findings were replicated in HeLa cells stably expressing Swedish mutated amyloid precursor protein (swAPP), with β-secretase inhibitor C3 treatment used as a negative control (SFigure 1).

**Figure 1.**
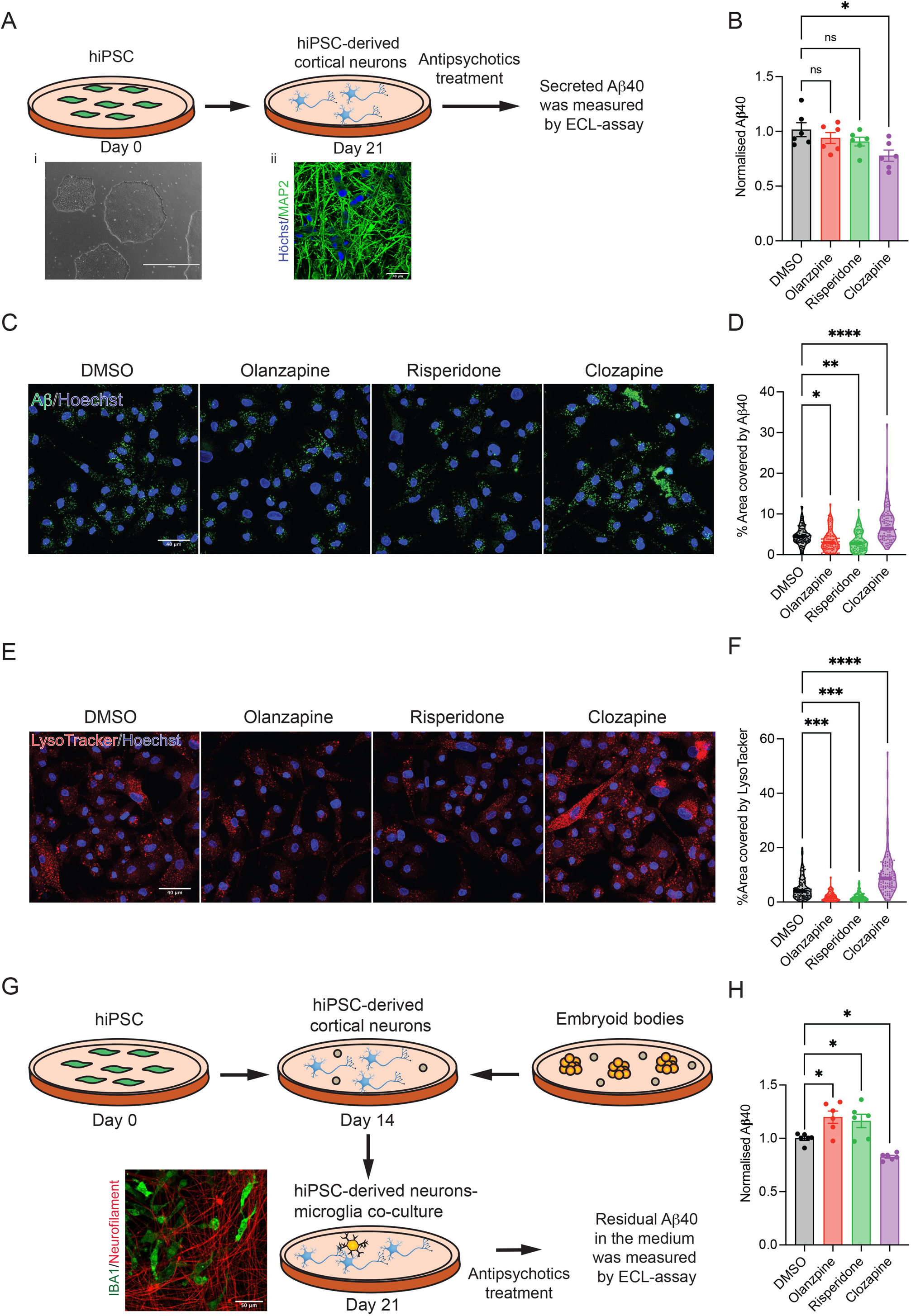
Antipsychotics alter Aβ secretion by neurons and Aβ clearance by microglia. **A.** Schematic representation of measurement of Aβ secretion by hiPSC-derived neurons. i. DIC image of iPSC culture, ii. Immunofluorescence image of iPSC-derived neuronal culture. MAP2 in green and nuclei were stained with Höchst (blue). **B.** hiPSC-derived cortical neurons were treated with Olanzapine (100 nM), Risperidone (100 nM) and Clozapine (1 μM) for 48h with a medium change in 24h. The medium is harvested to analyze the quantity of Aβ40 using electrochemiluminescence assay and the count normalized to total protein level in cell lysate. The error bar = SEM. Data was statistically analyzed via one-way ANOVA (Dunnet’s multiple comparisons test, *p<0.05, ns= not significant). **C-F.** iPSC- derived microglia were treated with Olanzapine (100 nM), Risperidone (100 nM) and Clozapine (1 μM) for 48h with a medium change in 24h. After treatment, cells were incubated with 250 nM Aβ40-HyLite-647 or 75 nM LysoTracker for 100 minutes. (**C**) Representative image of Aβ uptake assay. Aβ40 in green and nucleus were stained with Höchst (Blue). Scale bar is 40 μm. (**D**) Quantification of 1C. Data was statistically analyzed via one-way ANOVA (Dunnet’s multiple comparisons test, *p<0.05, **p<0.005, ****p<0.0001). n= 70-100 cells per conditions. (**E**) Representative image of LysoTracker (red). Nuclei were stained with Höchst (Blue). Scale bar is 40 μm. (**F**) Quantification of 1E. Data was statistically analyzed via one-way ANOVA (Dunnet’s multiple comparisons test, **p<0.005, ***p<0.0005, ****p<0.0001). n= 70-100 cells per conditions. **G.** Schematic diagram of preparation of neurons-microglia co-culture. In the representative image of co- culture, microglia were stained with anti-IBA1 antibody (green) and neurons were stained with anti-neurofliament antibody (red). **H**. Neuronal-microglia co-culture were treated with Olanzapine (100 nM), Risperidone (100 nM) and Clozapine (1 μM) for 48h with a medium exchange at 24h. The medium is harvested to analyze the quantity of Aβ using electrochemiluminescence assay. The error bar = SEM. Data was statistically analyzed via one-way ANOVA (Dunnet’s multiple comparisons test, *p<0.05).

### Antipsychotics modulate microglial-mediated Aβ clearance in a drug dependent manner

Total brain amyloid levels are controlled by the rate of both production and clearance. As the resident macrophage of the brain, microglia play a crucial role in the clearance of monomeric and fibrillar forms of Aβ (<u>Bolmont et al., 2008</u>, <u>Marzolo et al., 2000</u>). We therefore next examined the effect of Risperidone, Olanzapine and Clozapine on amyloid clearance by microglia. hiPSC-derived microglia monocultures (<u>Solomon et al., 2022</u>, <u>Couch et al., 2023</u>) were acutely exposed (48h) to the same concentrations of Risperidone, Olanzapine or Clozapine used in the neuronal studies and then incubated with fluorescently tagged Aβ40 (Figure 1C, SFigure 2). Acute Olanzapine and Risperidone exposure resulted in significant reductions in microglial uptake of Aβ40, whereas Clozapine significantly increased Aβ40 uptake by microglia (Figure 1C, D). These findings were consistent with changes in the level of acidic organelles of the cells, which are predominantly lysosomes, stained by LysoTracker DND-99 (Figure 1E, F). Similar findings were obtained using the BV2 immortalized mouse microglial cell-line (SFigure 3).

Taken together, these data suggest that acute Olanzapine and Risperidone exposure does not alter Aβ production by cortical neurons, but suppresses Aβ clearance by microglia, resulting in Aβ accumulation. Conversely, Clozapine treatment inhibits Aβ secretion by neurons yet facilitates Aβ clearance by microglia, leading to an overall reduction in Aβ levels. Since the properties of microglia are shaped by neuronal contact (<u>Haenseler et al., 2017</u>), to verify our monoculture findings, we acutely exposed hiPSC-derived neuronal-microglial co-cultures to the three antipsychotics, and measured residual Aβ levels in the medium (Figure 1G). In line with our monoculture findings, we observed a significant accumulation of Aβ in Olanzapine and Risperidone exposed cultures, while Clozapine led to a significant reduction in Aβ levels (Figure 1H).

### Clozapine treatment induces lysosomal biogenesis through TFEB

LysoTracker-DND-99 is a pH-sensitive dye, hence changes in staining with this marker could arise as a result of a change in lysosomal pH, a change in lysosomal levels, or both. The presence of basic tertiary amine groups in the three antipsychotics tested might be expected to increase lysosomal pH. However, lysosensor dextran yellow/blue did not show any significant alterations in lysosomal pH levels following acute exposure of microglia to the antipsychotics tested. By contrast, treatment with Baflomycin-A1, an inhibitor of vATPase, which pumps protons into lysosomes, led to an increase in lysosomal pH (Figure 2A). These data suggest alterations in LysoTracker levels observed in response to the antipsychotic treatments most likely reflect a change in lysosomal levels within the cell. We therefore quantified the levels of two lysosomal proteins, Lamp1 and Lamp2, and found that both were significantly increased in BV2 cells only following acute Clozapine exposure, but not after acute Olanzapine or Risperidone (Figure 2B-D). RT-qPCR analysis confirmed a clear induction of the expression of selected lysosomal genes (*Ctsd, Clcn7, Rab7*, etc.) in response to Clozapine exposure, but not olanzapine or risperidone (Figure 2E). Since this effect of Clozapine is present at the transcript level, we hypothesised Clozapine may be acting via TFEB (transcription factor EB) mediated CLEAR (coordinated lysosomal expression and regulation) network gene expression, resulting in lysosomal biogenesis (<u>Roczniak-Ferguson et al., 2012</u>, <u>Settembre et al., 2011</u>). To test this, we exposed HeLa cells stably expressing TFEB-GFP to Clozapine for 3h and observed an increase in TFEB nuclear translocation, leading to an increase in LysoTracker levels (Figure 2F-H). Collectively, these data suggest that Clozpaine induces TFEB-mediated lysosomal biogenesis, leading to increased phagocytic capacity.

**Figure 2.**
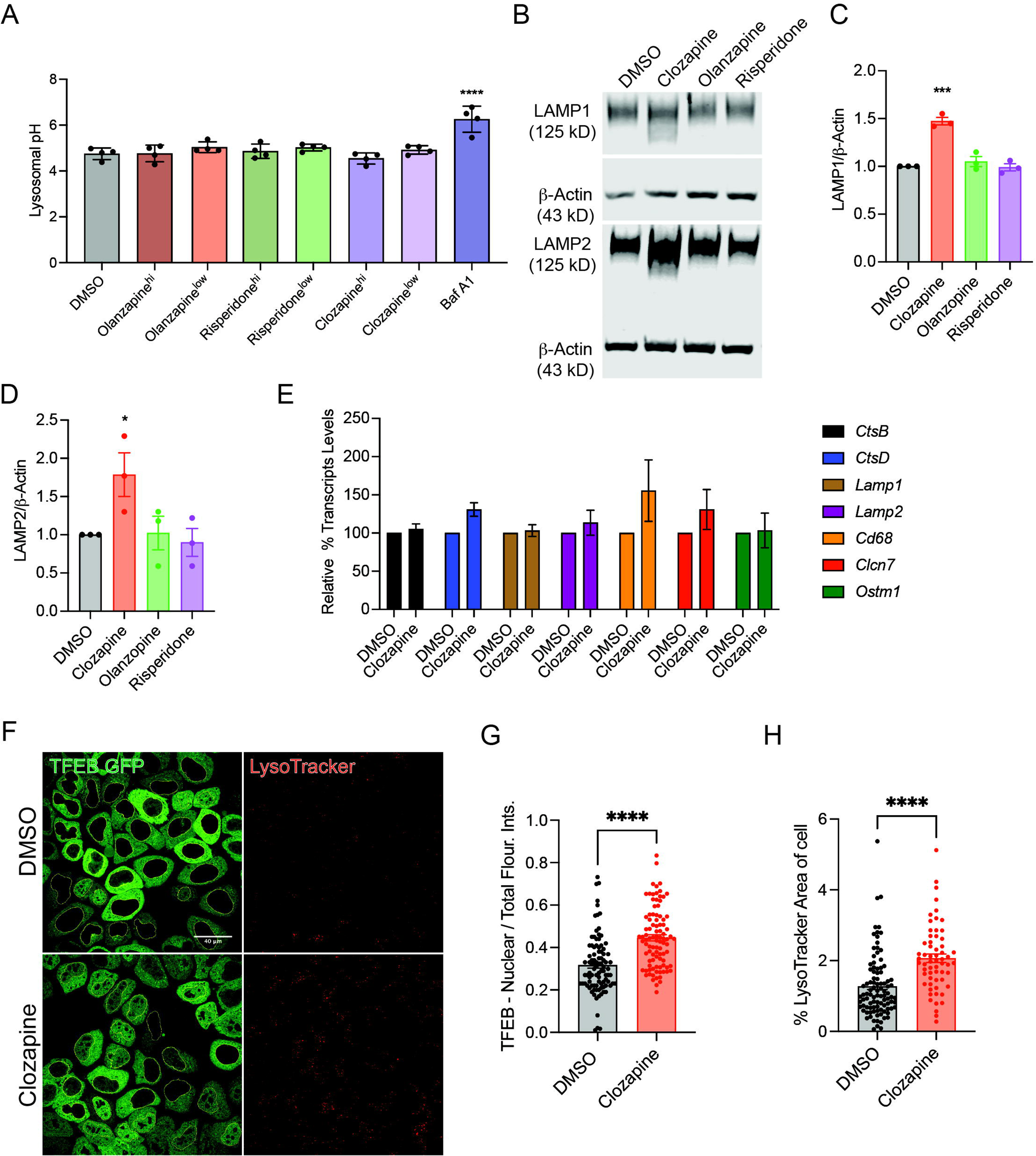
Clozapine increased lysosomal biogenesis through TFEB. **A.** BV2 cells were treated with two different doses of Olanzapine (100 nM, 10 μM), Risperidone (100 nM, 10 μM) and Clozapine (1μM, 25 μM) for 24 hours to assess changes in lysosomal pH using Lysosensor blue-yellow dextran. The error bar = SEM. Data was statistically analyzed via one-way ANOVA (Dunnet’s multiple comparisons test, ****p<0.0001). **B**. BV2 cells were treated with Olanzapine, Risperidone, Clozapine. Total protein was extracted and immunoblots were performed to probe LAMP1 and LAMP2. **C, D**. LAMP1 and LAMP2 immunoblots were quantified and normalized to β-Actin. Data was statistically analyzed via one-way ANOVA (Dunnet’s multiple comparisons test, *p<0.05, ***p<0.0005). **E.** BV2 cells were treated with Olanzapine, Risperidone and Clozapine for 6 hours. Total RNA was extracted to perform RT-qPCR. Targeted primers were used to probe lysosomal genes and data was analyzed using ΔΔCt method with normalization against β-Actin. **F.** HeLa TFEB- GFP were treated with Clozapine to analyze the nuclear-TFEB translocation. Representative images of GFP-TFEB (green) and LysoTracker (red). Scale bar = 40 μm. **G.** Integrated intensity of nucleus and the whole is quantified using ImageJ to find the TFEB translocation ratio. Quantification of %TFEB in the nucleus. **H.** Quantification %Area of the cell covered by LysoTracker. Data for G, H were statistically analyzed via non-parametric t-test (Mann- Whitney test, ****p<0.0001).

### Acute Clozapine exposure increases microglia mediated synapse engulfment

Our data thus far suggests that one potential cellular mechanism to explain the association of Risperidone and Olanzapine prescription with an increased risk for dementia is the reduced clearance of Aβ by microglia. Clozapine however, decreases Aβ production by neurons and increases its uptake by microglia, which is at odds with the observed increased risk for dementia following Clozapine exposure in our epidemiological data. In this context, it is important to note that in addition to Aβ accumulation, microglia also eliminate synaptic connections via multiple mechanisms during brain development and maturation (<u>Paolicelli et al., 2011</u>, <u>Weinhard et al., 2018</u>, <u>Eyo and Molofsky, 2023</u>). Reactivation of this developmental process likely contributes to the cognitive decline occuring in AD as a consequence of synapse loss (<u>Rajendran and Paolicelli, 2018</u>, <u>Hong et al., 2016</u>). We therefore next investigated the effect of the aforementioned antipsychotics on microglia-mediated engulfment of synaptic material. Using monocultures of hiPSC-derived microglia, we observed that acute Olanzapine and Risperidone exposure did not significantly alter synaptosome engulfment, however, acute Clozapine exposure markedly increased synaptosome engulfment (Figure 3A, B). These findings were replicated in BV2 cells (SFigure 4A-C).

**Figure 3.**
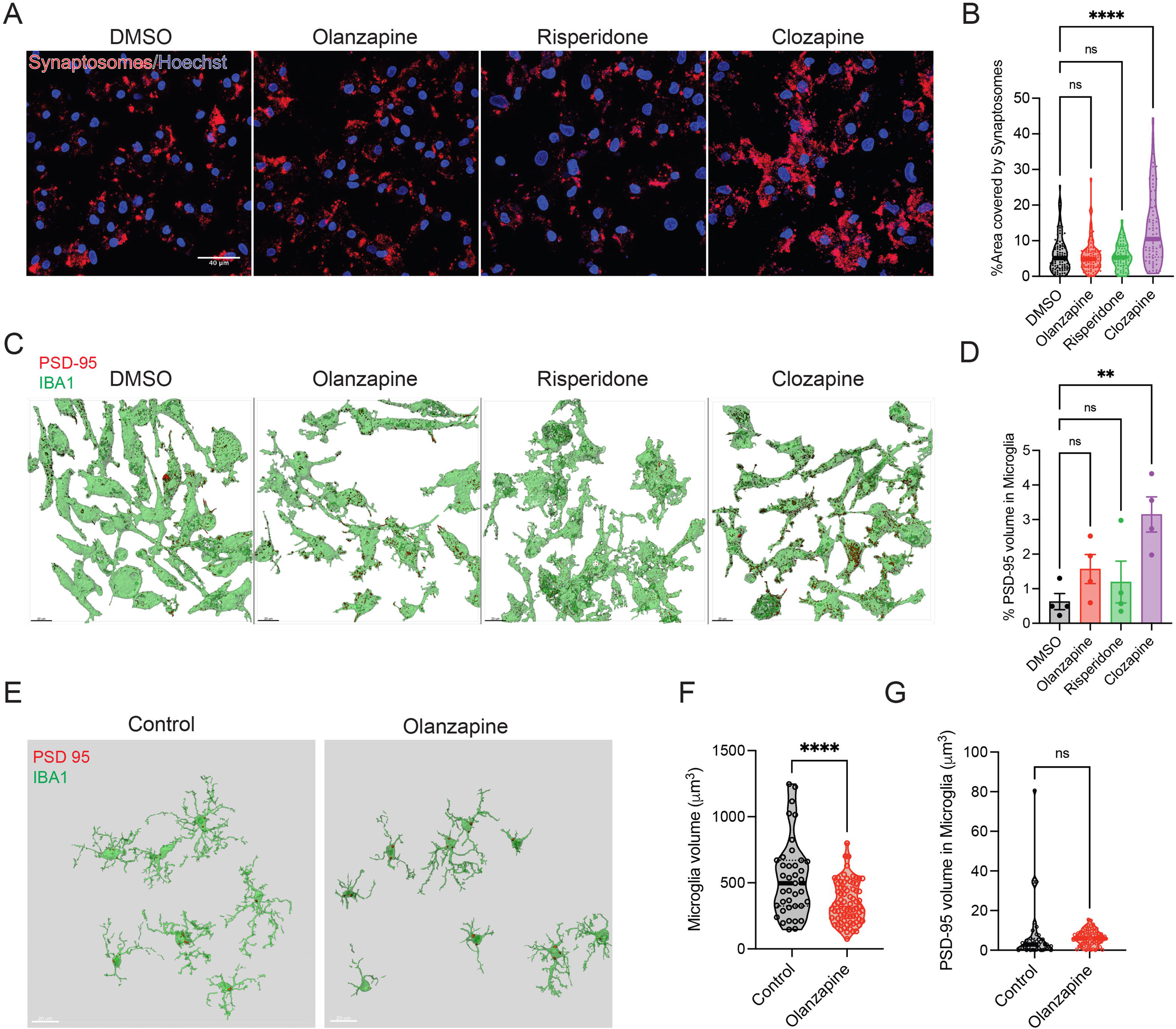
Antipsychotics modulate microglia mediated synaptosomes uptake: **A.** iPSC-derived microglia were treated with Olanzapine (100 nM), Risperidone (100 nM) and Clozapine (1 μM) for 48h with a medium exchange at 24h. After treatment, cells were incubated with ^td^Tomato-tagged synaptosomes for 100 minutes, fixed with PFA 4%, imaged with Nikon A1R. Representative images of Synaptosomes (red) upatake). Nuclei were stained with Höchst (Blue). Scale bar is 40 μm. **B.** Quantification of 3A. Data was statistically analyzed via one-way ANOVA (Dunnet’s multiple comparisons test, ns= not significant, ****p<0.0001). n= 70-100 cells per conditions. **C.** Neuronal-microglia co-culture were treated with Olanzapine (100 nM), Risperidone (100 nM) and Clozapine (1 μM) for 48h with single medium change at 24h. Cells were stained with Iba1 (green) and PSD-95 (red). Image shows 3D-reconstruction of microglia and internalized PSD-95 volume. **D**. Quantification of 3C. Data was statistically analyzed via one-way ANOVA (Dunnet’s multiple comparisons test, ns= not significant, **p<0.005). **E-F** Synaptic pruning was analyzed in somatosensory-motor cortex of naïve male rat brains. (E) Representative images. Iba1 (green), PSD95 (red). Scale bar= 20 μm. (F) Quantification of microglia volume (3E). **G.** Quantification of internalized PSD95 volume (3E). Data for F, G were statistically analyzed via non-parametric t-test (Mann-Whitney test, ns= not significant, ****p<0.0001). n=40-70 cells per condition.

As with Aβ uptake, we validated these findings using hiPSC-derived neuronal-microglial co-cultures. Cells were acutely exposed (48h) to each antipsychotic or vehicle and synapse engulfment was analyzed by measuring the amount of post-synaptic density protein-95 (PSD-95) inside microglia. Clozapine exposure significantly increased the volume of PSD-95 identified within IBA1+ microglia, while Risperidone and Olanzipine exposure had no significant effect (Figure 3C, D). These data provide initial evidence that acute exposure to Clozapine, but not Risperidone or Olanzapine, could enhace microglia-mediated synaptic engulfment, under the conditions tested.

To complement our *in vitro* data, we examined the effect of *chronic* Olanzapine treatment on PSD-95 engulfment by microglia *in vivo*. Naïve male Sprague-Dawley rats were exposed to Olanzapine using osmotic mini pumps for 28 days (7.5 mg/kg/d; n=4) or a common drug vehicle (n=4) as previously reported (Onwordi et al. 2020; Halff et al., 2021).

This exposure resulted in a terminal drug plasma concentration of 16.3-26.9 ng/mL (Onwordi et al. 2020; Halff et al., 2021). Using immunostaining and confocal microscopy we examined the morphology of Iba1+ microglia and the volume of PSD-95 engulfment by Iba1+ microglia in the rat somatosensory cortex. This region was selected on the basis of prior work suggesting that chronic Olanzapine treatment causes an increase Iba1+ microglia density and amoeboid morphology in this region of the rat brain (<u>Cotel et al., 2015</u>). We observed that Iba1+ microglia from Olanzapine-treated rats were less ramified and had reduced volume, compared with vehicle-treated controls (Figure 3E, F), consistent with and confirming the aforementioned observations (<u>Cotel et al., 2015</u>). There was however no statistically significant increase in the volume of PSD-95 within the Iba1+ microglia when comparing Olanzapine- and vehicle-exposed rats (Figure 3G). These data are consistent with our *in vitro* observations in human iPSC neuron-microglia co-cultures and prior data which observed no effect of either haloperidol or olanzapine on specific pre- and post-synaptic proteins in the rat brain (Onwordi et al. 2020; Halff et al. 2021).

## Discussion

Several epidemiological studies provide evidence for an association between the a diagnosis of schizophrenia and an increased risk for dementia (<u>Coupland et al., 2019</u>, <u>Stroup et al., 2021</u>, <u>Tampi et al., 2016</u>, <u>Vigen et al., 2011</u>). To what extent antipsychotic drugs may mediate this association is however unclear. Our pharmaco-epidemiological study of the Swedish prescription database suggests that prescription of Olanzapine, Risperidone, and Clozapine is associated with increased risk of not only AD but also other forms of dementia including vascular dementia. The association was noted among males and females and across all age groups and across different psychiatric diagnoses for which these drugs may be prescribed.

To begin to understand the cause of this association, we sought to explore the cellular processes that may underpin our epidemiological findings. Specifically, we investigated the acute effects of these antipsychotics on Aβ production and clearance, and lysosomal levels using methods previouslty reported by our group (<u>Mondal et al., 2020</u>). Neither Olanzapine nor Risperidone altered Aβ secretion in human iPSC-derived cortical neurons, but reduced microglial Aβ uptake, which was positively correlated with microglia LysoTracker Red levels. This suggests that Olanzapine and Risperidone may lead to Aβ accumulation, reducing clearance of the peptide, a finding confirmed in our neuronal-microglial co-culture study. Conversely, Clozapine treatment significantly reduced Aβ load by reducing neuronal production and increasing microglial phagocytosis.

This Clozapine indued reduction in Aβ load was surprising given that this antipsychotic demonstrated the largest increased risk for AD in our epidemiological dataset. Logically these data suggests that Clozapine acts via a different mechanism to mediate this increased dementia risk. Consistent with this view, we found that while Olanzapine and Risperidone treatment had no impact on microglial-engufment of synaptosomes or PSD-95, Clozapine treatment resulted in a significant increase in synaptosome uptake. The absence of any effect of Olanzapine on synapse engulfment, either *in vitro* or *in vivo*, is consistent with recently published studies demonstrating that chronic Olanzapine exposure was not associated with a reduction in synaptic vesicle glycoprotein 2A (SV2A) protein levels, or the pan-post-synaptic marker Neuroligin (I-IV) in the naive male rat brain (<u>Halff et al., 2020</u>, <u>Onwordi et al., 2020</u>). These data support the notion that Olanzapine is unlikely to contribute to synaptic loss in schizophrenia as measured *in vivo* using SV2A selective positron emission tomography (PET) (<u>Onwordi et al., 2020</u>). Of note, structural magnetic resonance imaging (sMRI) studies of individuals with treatment-resistant schizophrenia, provide evidence for cortical thinning and sub-cortical volume reductions relative to age-matched healthy controls after switching to Clozapine treatment (<u>Ahmed et al., 2015</u>, <u>Krajner et al., 2022</u>). Moreover, the cellular basis of such tissue loss cannot be established from these studies. However, electron microscopy analysis of rat cortical brain tissue following long-term Clozapine exposure provides evidence of a relative shift in the distribution of excitatory and inhibitory elements synapsing onto dendritic shafts and, to a lesser extent, spines in layer VI of the rat medial prefrontal cortex, suggestive of some synaptic rearrangements (<u>Vincent et al., 1991</u>). In the present study, we were unable to test the effect of chronic Clozapine exposure on synaptic engulfment by microglia *in vivo* due to solubility issues that precluded the use of clinically-comparable dosing via osmotic minipumps in rats as reported by Kapur and colleages (<u>Kapur et al., 2003</u>). Overall, further research is urgently required to confirm *in vivo* the effects of Clozapine on synaptic density and how this relates to cognition and risk for dementia.

A large population-based cohort study demonstrated that individuals with schizophrenia had a two-fold increase in the risk for developing dementia (<u>Ribe et al., 2015</u>). Post-mortem studies show a reduction in dendritic spine density and synaptic markers in the brains of schizophrenia patients relative to age-matched controls (<u>Green, 2006</u>, <u>Glantz and Lewis, 2000</u>, <u>Osimo et al., 2019</u>). As mentioned above, a recent PET study has shown a reduction in SV2A radioligand binding in chronic schizophrenia patients (<u>Onwordi et al., 2020</u>) but not in the early stages of the illness (Onwordi et al Biological Psychiatry, 2023). Furthermore, a schizophrenia patient-derived cell model displayed higher synapse elimination by microglia than cells derived from healthy controls (<u>Sellgren et al., 2019</u>). Collectively, these studies suggest that schizophrenia *per se* is associated with alterations in microglial function and synaptic pruning, which, speculatively, may be further exacerbated by treatment with Clozapine. Reductions in synapses may at least partly underlie the increased risk of dementia development in schizophrenia patients, and alterations in Aβ clearance or synaptic pruning induced by Olanzapine, Risperidone or Clozapine, as demonstrated in this report, may synergistically interact with these illness related factors to further increase risk for dementia. In human studies, it is difficult to completely isolate the effect of the underlying disease (i.e., schizophrenia or bipolar disorders) from the effect of the antipsychotics, or vice versa. In our analysis, we separately analyzed individuals with a diagnosis of schizophrenia or bipolar disorders and individuals without such diagnoses. The fact that positive associations were noted in both analyses suggests that the use of antipsychotics might indeed be associated with an increased risk of dementia, regardless of the underlying psychiatric disorder. A view that is reinforced by our findings that these drugs impacted on key processes implicated in AD in our healthy control cells.

While the majority of our cellular studies were focused on understanding the mechanisms by which different antipsychotics may increase the risk for AD, alterations in microglial phagocytosis likely has implications for other dementias, consistent with our epidemiological findings. In support of this, high doses of atypical antipsychotics are associated with increased risk and poorer survival outcomes in parkinsonism (<u>Rochon et al., 2005</u>) (<u>Marras et al., 2012</u>), providing broader evidence for a link between neurodegeneration and antipsychotic treatments that may, based on our current findings, at least in part arise as a result of altered microglial function. However, atypical antipsychotics have also been associated with an increased risk of myocardial infarction and stroke (<u>Brauer et al., 2015</u>, <u>Douglas and Smeeth, 2008</u>, <u>Hsu et al., 2017</u>), a risk that is further increased in patients with dementia (<u>Douglas and Smeeth, 2008</u>). This could be indicative of a broader impact of antipsychotics on metabolic and vascular function, and may provide an alternative mechanism for how antipsychotics increase the risk of vascular dementia as observed in our epidemiological study. Clearly further experimentation is required to fully understand the relative contributions of altered microglial cell function and vascular changes to the increased risk of neurodegenerative disease and dementia associated with antipsychotic use.

A growing body of evidence demonstrates possible side effects of antipsychotics, including the increased risk of mortality in patients with dementia as well as in individuals without dementia (<u>Ralph and Espinet, 2018</u>). Therefore, an increased risk of dementia is concerning in the context of antipsychotic use. It is important to note however, that the studied antipsychotics are generally efficacious and well-tolerated, and their use is clinically very important to prevent relapse, hospitalisation, and death (<u>Huhn et al., 2020</u>). The celluar effects we observed were also obtained using healthy donor cell lines. Hence there may be drug-by-disease interactions we have not captured. It will therefore be important to study these drug effects using patient-derived lines either from individuals with schizophrenia, or in the context of genetic risk for AD that are known to influence microglia form and function. It is also important to note that although our epidemiological study examined the role of antipsychotic use up to nine years before a first specialist visit for dementia, our functional experiments were conducted under acute conditions in a cellular system. Caution must therefore be taken before applying these findings to the broader context of long-term chronic drug use in human patients. Nonetheless, in the current study, we have identified a distinct cellular mechanism that may explain this risk. Understanding the differential impacts of these drugs on the various aspects of dementia pathology will be crucial in helping to inform our approaches to managing patient treatment regimes in the future. The data presented in this report also provide key evidence supporting the importance of microglial cell function in the onset and progression of AD.

## Materials and Methods

### Register-based pharmacoepidemiological study

The Swedish Prescribed Drug Register encompasses data on all prescribed and dispensed medications since July 2005 in Sweden, including information on the Anatomical Therapeutic Chemical (ATC) codes, dosage, and dates of prescription and dispense (<u>Wallerstedt et al., 2016</u>, <u>Wettermark et al., 2008</u>).

We included all individuals in the Swedish Total Population Register that were born in Sweden and living in Sweden on July 1, 2005 as the study population. We followed these individuals from July 1, 2005 until their diagnosis of dementia, emigration out of Sweden, death, or December 31, 2013, whichever came first. The individual follow-up was performed through cross-linking the Total Population Register to the Swedish Patient Register, the Migration Register, and the Causes of Death Register using the individually unique personal identity numbers. Through the Patient Register, which includes nationwide information on all hospital discharge records from 1987 onward and >80% of outpatient specialist care in Sweden from 2001 onward (<u>Ludvigsson et al., 2011</u>), we identified individuals with a newly diagnosed dementia during follow-up, according to the 10th Swedish revision of the International Classification of Disease codes (F00, G30 for AD, F01 for vascular dementia, and F02, F03, G311, G318A, F051 for other dementias). The date of the first hospital visit concerning dementia was used as the date of diagnosis. The accuracy of dementia definition based on the Patient Register is satisfactory (positive predictive value: 81%) (<u>Rizzuto et al., 2018</u>).

We conducted a nested case-control study within the above study base to assess the association of antipsychotic use with the future risk of dementia. Using the method of incidence density sampling, we randomly selected up to five controls per dementia case who were individually matched to the case by sex, year of birth, and area of residence. Eligible controls were individuals who were alive and free of dementia diagnosis on the diagnosis date of their corresponding cases. The date of diagnosis and date of selection were used as the index date for cases and controls, respectively. We identified 85,721 patients with a newly diagnosed dementia during follow-up (including 25,241 cases of AD, 14,081 cases of vascular dementia, and 46,399 cases of other dementias), and selected 426,599 matched controls for these cases. The group with other dementias was likely a mixture of patients with AD, vascular dementia, and other dementias.

Through linking the case-control study to the Prescribed Drug Register, we ascertained information on all dispensed Olanzapine, Risperidone and Clozapine from July 1, 2005 until the index date of both cases and controls, using the ATC codes N05AH03, N05AX08 and N05AH02 respectively. Antipsychotic use was defined first as a binary variable (use or no use) and then as a categorical variable according to the tertile distribution of cumulative defined daily doses (cDDDs). The cDDDs were calculated as the total dispensed number of defined daily doses between July 1^st^, 2005 and the index date. We used conditional logistic regression models to estimate odds ratios (ORs) and their 95% confidence intervals (CIs) of dementia in relation to use of the three antipsychotics. To assess the temporal pattern of the studied associations, we estimated the ORs for antipsychotics used during the 0-1, 1-2, 2-3, 3-4, 4-5 and 5-9 years before the index date. To alleviate the potential concern of reverse causation (i.e., some symptoms of dementia might lead to a misdiagnosis of psychiatric disorders and the resultant use of antipsychotics), we studied only antipsychotics used more than 3 years before the index date in the main analysis. Two sensitivity analyses were also performed by studying only antipsychotics more than 4 or 5 years prior to the index date to validate the robustness of results from the main analysis. To assess the potential effect modification by age and sex, we also stratified the main analyses by age at index date (<70 and ≥70 years) and sex (male or female). In addition to any dementia, we also studied AD, vascular dementia and other dementias separately. Finally, to alleviate the concern of indication bias, i.e., the association between use of antipsychotics and risk of dementia might merely represent the association between the indicating conditions of the antipsychotics (e.g., schizophrenia or bipolar disorders) and dementia, we performed a stratified analysis by separately analysing individuals with a diagnosis of schizophrenia or bipolar disorders and individuals without such diagnosis. History of schizophrenia and bipolar disorders was ascertained from 1969 until index date from the Swedish Patient Register. Data analyses were performed using SAS version 9.4 (SAS Institute Inc, Cary, NC). A two-sided P<0.05 was considered statistically significant for the primary exposure. The analysis was approved by the Regional Ethical Review Board in Stockholm, Sweden.

### Cell lines and compounds

BV2 cells were cultured and maintained at 37°C in low glucose DMEM (11885084, ThermoFisher) supplemented with 10% FCS (10091148, ThermoFisher) and 1% Pen/Strep (15140122, ThermoFisher). HeLa swAPP (HSA) cells were similarly maintained in low glucose DMEM with 10% FCS and 1% Pen/Strep with the addition of 0.1% geneticin (10131035, ThermoFisher) (50 mg/ml stock) and 0.1% zeocin (R25001, ThermoFisher) (100 mg/ml stock). HeLa TFEB-GFP were similarly maintained in low glucose DMEM with 10% FCS and 1% Pen/Strep with the addition of 0.1% geneticin (10131035, ThermoFisher) (50 mg/ml stock). Olanzapine (Sigma-Aldrich, 01141), Risperidone (Sigma-Aldrich, R3030), Clozapine (Sigma-Aldrich, C6305) were diluted in DMSO and DMSO was used as vehicle control.

### Generation of human iPSC derived cortical neurons

Cortical neurons were differentiated by adapting the Fernandopulle et al. protocol (<u>Fernandopulle et al., 2018</u>). The iPSC cells were cultured on Geltrex coated (1:100 in DMEM/F12), in E8-Flex medium (A2858501, ThermoFisher). Cells were transduced using CRISPR-cas9 RNP to stably integrate a differentiation construct, doxycycline inducible NGN2 expression cassette, into the AAVS1 safe harbour driving the cellular differentiation into cortical neurons. NGN2 stably expressing iPSCs were then induced in DMEM/F12 with HEPES (11330032, ThermoFisher) supplemented with; N2 (5 ml per 500 ml) (17502048,

ThermoFisher), NEAA (5 ml per 500 ml) (11140050, ThermoFisher), Glutamax (5 ml per 500 ml) (35050038, ThermoFisher) and doxycycline (2μg/ml) (D9891, Sigma). After 3 days, the cells were transferred to Poly-D-Lysine (PDL) and Laminin (1:100 dilution in PBS) (23017015, ThermoFisher) coated plates and culture in Neurobasal media (05790, STEMCELL Technologies) supplemented with; 1X B27 (17504044, ThermoFisher), 10 ug/ml BDNF (450-02, PeproTech), 10 ug/ml NT-3 (450-03, PeproTech) and Laminin (50 ul of 1 mg/ml per 50 ml) for up to 12 days.

### Antipsychotic treatment to iPSC-derived neurons

iPSC-derived cortical neurons were further maintained in neuronal maintenance medium (as listed in the supplementary method) until 21 days. Following this, neurons were treated with DMSO and antipsychotics (Olanzapine: 100 nM, Risperidone: 100 nM and Clozapine: 1 μM) for 48h with one medium change at 24h. The serected Aβ was measured using electrochemiluminescence assay and normalized to total protein level.

### Measurement of Aβ using Electrochemiluminescence assay (Meso Scale discovery)

Secreted Aβ40 level in the medium was measured by Electrochemiluminescence assay (MSD) using supplied protocol. Each well of the pre-coated assay plate was first blocked using 150 μl diluent-35, then incubated with 25 μl of the conditioned medium along with detection antibody (6E10, MSD). Wells were washed 3 times with 150 μl Tris-wash buffer and detection was performed using 150 μl of 2x Read buffer.

### iPSC generation of microglia

Macrophages and macrophage-derived microglia were generated following Haenseler et al. (<u>Haenseler et al., 2017</u>). In brief, embryonic bodies (EBs) were generated from iPSCs by plating on an AggreWell 800 (34850, StemCell Technologies) in E8-Flex medium supplemented with 50 ng/ml BMP4 (120-05ET, Peprotech), 50 ng/ml VEGF (PHC9394, ThermoFisher) and 20 ng/ml SCF (300-07, PeproTech) for 3 days. EBs were transferred onto a T75 flask and maintained in X-VIVO15 (BE02-060F, Lonza) medium supplemented with 100 ng/ml M-CSF (300-25, PeproTech), 25 ng/ml IL-3 (PHC0031, ThermoFisher), 2 mM Glutamax (35050061, ThermoFisher) and 0.055 mM β-mercaptoethanol (31350-010, ThermoFisher). After 4 weeks, precursor cells started to emerge. The precursor cells were collected and differentiated to microlgia over 7-days in neuronal maintenance medium supplemented with 100 ng/ml M-CSF, 100 ng/ml IL-34 and 10 ng/ml GM-CSF.

### Antipsychotic treatment to iPSC-derived microglia

50,000 microglia precoursor cells were plated in each well of an 8 well chamber slide (80827, Ibidi) and differentiated to microglia according to above mentioned protocol. After 7 days, iPSC-derived microglia were treated with DMSO and antipsychotics (Olanzapine: 100 nM, Risperidone: 100 nM and Clozapine: 1 μM) for 48h with one medium change in 24h. The cell were then incubated with 75 NM LysoTracker DND-99 (L7528, ThermoFisher), 250 nM Aβ40 HyLite Fluor 657 (AS-60493, AnaSpec) or ^td^ purified from mouse brain for 100 min. Cells were washed with PBS and then fixed using 4% PFA for 30 minutes. Nucleus were stained with Hoechst (62249, ThermoFisher). Plates were then imaged using a A1R inverted confocal microscope.

### Antipsychotic treatment to iPSC-derived neuronal-microglial co-culture

For iPSC-derived neuronal-microglial co-culture, iPSC-derived neurons were plated and maintained in 48 well plate (75,000 in each well) or 8 well chamber slide (60,000 in each well) until day-12. Then, iPSC derived microglial precursor cells were plated on it in one third number to neurons. The co-culture were further maintained for 7 days. Following this, iPSC-derived neurons-microglia co-culture were treated with DMSO and antipsychotics (Olanzapine: 100 nM, Risperidone: 100 nM and Clozapine: 1 μM) for 48h with one medium change in 24h. For 48 well plates, the serected Aβ were were measured using electrochemiluminescence assay. For 8 well chamber slides, cells were washed with PBS and then fixed using 4% PFA for 20 minutes. Cells were stained with IBA1 (1:500 dilution; Wako 019-19741) and PSD-95 (1:100; ThermoFisher MA1-045). Nucleus were stained with Hoechst (62249, ThermoFisher). Plates were then imaged using a A1R inverted confocal microscope. The PSD-95 voulme inside microglia (IBA1 volume) were measured using 3D reconstruction technique by IMARIS.

### Synaptosomes preparation

B6.Cg-Tg(Camk2a-cre)T29-1Stl mouse was crossed with B6.Cg-Gt(ROSA)26Sortm14(CAG-tdTomato)Hze mouse. 2 month old pups were then sacrificed using intracardiac perfusion, after deep anesthesia. The brains tissue was the homogenized in Syn-PER Reagent (Thermo Fisher) containing protease inhibitor on ice and synaptosomes were isolated according to the provided protocol. Briefly, first homogenate was centrifuged at 1200 × g for 10 minutes at 4°C. pellet was discarded and supernatant was further centrifuged at 15,000 × g for 20 minutes at 4°C. The synaptosomes pellet was suspended in Syn-PER reagent containing 5% DMSO (500μL for 400mg of brain tissue). The synaptosomes were aliquoted and stored -20°C for further use.

### Phagocytic Imaging assay of BV2 cells

An 8-well 1.5h imaging plate (80827, Ibidi) was coated with PDL and seeded with BV2 cells at a density of 7,000 cell per well. The cells were left for 36 hours and then treated with DMSO or antipsychotics (Olanzapine: 10 μM, Risperidone: 1 μM and Clozapine: 25 μM) for 24h. The cell were then incubated with 75 NM LysoTracker DND-99 (L7528, ThermoFisher), 250 nM Aβ40 HyLite Fluor 657 (AS-60493, AnaSpec) or ^td^Tomato-tagged synaptosomes from mouse brain for 1h. Cells washed with PBS and then fixed using 4% PFA for 30 minutes. Nucleus were stained with Hoechst (62249, ThermoFisher). Plates were then imaged using a A1R inverted confocal microscope.

### Aβ Phagocytosis assay in BV2 cells

BV2 cells were seeded in a PDL coated 96-well plate at a density of 7,000 cell per well. The cells were left for 36 hours and then treated with DMSO or antipsychotics (Olanzapine: 10 μM, Risperidone: 1 μM and Clozapine: 25 μM) for 10h. The cell were then incubated with HSA conditioned medium containg Aβ for 14h along with antipsychotics treatment. Medium were collected and residual Aβ40 was measured using Electrochemiluminescence assay. The uptaken Aβ40 was then calculated according to the initial Aβ counts. Aβ40 count were normalized to total protein amount in cell lysate.

### Amyloid production assay on HeLa swAPP cells

HeLa swAPP cells were plated at a density of 5,000 onto 96-well plates. After 36h, cells were incubated with medium supplemented with either DMSO or antipsychotics (Olanzapine: 10 μM, Risperidone: 1 μM and Clozapine: 25 μM) for 24h. Medium were changed with fresh medium and collected for 4h. Secreted Aβ40 was measured using Electrochemiluminescence assay and normalized to protein concentration in cell lysate. 5 μM C3 treatment was used as control.

### Lysosensor assay

For detection of pH changes, BV2 cells were treated with Olanzapine (high: 10 μM and low: 0.1 μM), Clozapine (high:25 μM and low:1 μM) and Risperidone (high:1μM and low:0.1 μM), 100 nM Bafilomycin A1 or DMSO. Cells were futher incubated with lysosensor dextran yellow/blue (500 μg/mL) (L22460, Thermo) 23 hours. 10 μM monesin (M5273, Sigma) treatment was used for control curve and the medium was replaced with buffer solution made up 5 mM NaCl, 115 mM KCl, 1.2 mM MgSO4 and 25 mM 4-morpholineethanesulfonic acid (MES) (M1317, Thermo) titrated between pH 4.5 and 7.0 for 10 minutes. For the experimental condition cells were washed once with PBS and incubated with PBS before being read twice at 335:425 and 381:521 on a Spectofluroescent plate reader. The ratio between 425 and 521 was determined and using the developed standard curve was read off to give an estimated lysosomal pH.

### Immunoblotting

For detection of intracellular proteins, whole cell extracts were prepared using a lysis buffer (1%NP40 and 0.1% SDS) supplemented with proteinase inhibitors. Extracts were subjected to SDS-PAGE using pre-cast gels (Invitrogen). Proteins were transferred onto membranes, and blocked with PBS containing 3% (w/v) BSA. The membranes were incubated with primary antibodies (LAMP1 (1:1000; DSHB 1D4B), LAMP2 (1:1000; DSHB ABL-93), β-Actin (1:10000; Abbcam ab8226)), followed by secondary probing with Licor fluorescent antibodies for 1h at room temperature. Immunoblotted proteins were detected using an Odyssey CLx Imager.

### RT-qPCR

Total RNA was isolated using DIRECT-ZOL (Zymo research). RNA purity (A260/A280, A260/A230) and concentration were measured using a Nanodrop spectrophotometer. 500 ng of total RNA was used for reverse transcription using super script IV one-step reverse transcription kit (Bio-Rad). RT-PCR analysis was performed using Taq man probes following manufacturer’s instructions using a 7900HT Fast Real-Time PCR system (Applied Biosystems). Assays were performed in triplicate and expression levels of genes were normalized against β-Actin using the ΔΔCt method.

### Animal model for chronic Olanzapine exposure

This study using archival tissue from a previous study (Halff et al. 2021) and no new animals were used. All animal experiments were carried out in accordance with the Home Office Animals (Scientific Procedures) Act (1986) and European Union (EU) Directive 2010/63/EU, with the approval of the local Animal Welfare and Ethical Review Body (AWERB) panel at King’s College London (KCL). Male Sprague-Dawley rats (Charles River UK Ltd, Margate, UK), initial body weight 220−270 gram (6− 10 weeks of age) were housed four per cage in conventional plastic cages (38 × 59 × 24 cm, Tecniplast, UK) containing sawdust, paper sizzle nest and cardboard tunnels. Animals were maintained under a 12 -h light/dark cycle (07.00 lights on) with food and water available *ad libitum*. Room temperature and humidity were maintained at 21 ± 2 11C and 55 ± 5%, respectively. Animals were habituated for a minimum of 7 days before experimental procedures. Animals received continuous administration of either common vehicle (n=4; β-hydroxypropylcyclodextrin, 20% wt/vol, acidified by ascorbic acid to pH 6) or OLZ (n=4; 7.5 mg/kg/ day; Sigma-Aldrich, Gillingham, UK) for 28 days, via osmotic minipumps (Alzet, Model 2ML4), inserted subcutaneously on the back flank, as previously described elsewhere (<u>Halff et al., 2021</u>, <u>Peris-Yague et al., 2020</u>). Blood plasma levels achieved using these doses and delivery by osmotic pumps have previously been shown to be consistent with clinically comparable dopamine D2 receptor (D2R) occupancy and plasma levels (<u>Kapur et al., 2003</u>, <u>Vernon et al., 2011</u>). Minipumps filled with Olanzapine or vehicle solutions were inserted subcutaneously on the back flank under isoflurane anaesthesia (5% induction, 1.5 % maintenance delivered in an 80/20 % medical air/oxygen mix). Weight was monitored daily in the week following surgery, and twice a week in the following weeks. After 28 days of administration, animals were terminally anaesthetised by injection of sodium pentobarbital (60 mg/kg, intraperitoneal) and culled by cardiac perfusion using heparinised (12.5 U/mL) ice-cold phosphate buffered saline (PBS). A blood sample was collected for estimation of drug levels done commercially using tandem mass spectrometry (Cyprotex, Macclesfield, UK). Brain tissues were rapidly extracted on a chilled plate, hemi-sected and post-fixed overnight in 4% paraformaldehyde (PFA) at 4 11C. Brain hemispheres were then washed once in PBS, incubated in PBS-buffered 30 % sucrose solution for 48 h at 4 11C, and then snap-frozen on dry ice and stored at − 70 11C. The left hemisphere of each animal was serially sectioned (30 μm-thick coronal sections, interval 1/12, 360 μm spacing between sections within a series) on a cryostat at − 18 11C and stored in tissue cryoprotection solution (25 % glycerol, 30 % ethylene glycol, 45 % 1x PBS pH 7.4, 0.05 % sodium azide) at − 20 11C until further processing for immunostaining.

### Immunostaining

For antibody staining, brain sections were permeabilized at room temperature in PBS containing 0.5% Triton X-100 (Sigma) for 10 min and followed by blocked using blocking solution (PBS, 2% BSA, 0.5% Triton X-100) for 1h in RT. Sections were incubated overnight with anti-Iba1 (1:500 dilution; Wako 019-19741) and anti-PSD-95 (1:100; Millipore MAB1596) antibody at 4°C. Brain sections were then washed with PBS containing 0.5% Triton X-100 and were incubated 2 hr RT with Alexa-fluorophore-conjugated secondary antibodies (Invitrogen). Nucleus were stained with Hoechst (62249, ThermoFisher). Plates were then imaged using a A1R inverted confocal microscope. The PSD95 voulme inside microglia (Iba1 volume) were measured using 3D reconstruction technique by IMARIS.

## Authors’ Contributions

MM planed the experiments. MM, SS, NKS, IC, EFH, MCC, PRM and AV performed the experiments. JS, FF performed the epidemiological study. MM wrote the first draft of the manuscript. All authors reviewed and edited the manuscript.

## Data Availability

All data produced in the present work are contained in the manuscript.

## Acknowledgements

We thank the Wohl Cellular Imaging Centre at King’s College London for help with confocal microscopy. We thank Dr. S. Ferguson for the generous support with TFEB-GFP cell lines. We also acknowledge the funding of the Dementia Research Institute, Van Geest Trust, Swedish Research Council (grant No. 2019-01088), Alzheimer’s Research UK (RE18385), and Karolinska Institutet (Senior Researcher Award and Strategic Research Area in Epidemiology). We would like to thank Els F. Halff for her help with animal experimentation. ACV acknowledges financial support for this study from the Medical Research Council (New Investigator Research Grant (NIRG), MR/N025377/1). The work (at King’s College, London) was also supported by the Medical Research Council (MRC) Centre grant (MR/N026063/1).

## Figure Legends

**SFigure 1. Clozapine reduces secreted A**β **levels in HeLa swAPP cells.** HeLa swAPP cells were treated with Olanzapine (10μM), Risperidone (1 μM) and Clozapine (25 μM) for 24 hours and the medium were collected for 4h to analyze the quantity of Aβ using electrochemiluminescence assay (MSD) and the count normalized to total protein level in cell lysate. DMSO was used as control. The error bar = SEM. Data was statistically analyzed via one-way ANOVA (Dunnet’s multiple comparisons test, ns= not significant, ****p<0.0001).

**SFigure 2.** Schematic diagram of hiPSC derived microglia preparation and experimental plan.

**SFigure 3. Antipsychotics alter microglia mediated A**β**40 uptake. A.** Experimental overview of Aβ uptake assay. **B.** BV2 microglia cells were treated with DMSO or antipsychotics (Olanzapine: 10 μM, Risperidone: 1 μM and Clozapine: 25 μM) for 10h followed by incubated overnight with supernatant taken from HeLa (swAPP) containing Aβ. The residual Aβ was quantified using MSD and normalized to total protein. Uptake Aβ was quantified through calculation the change from initial, control loaded Aβ amounts. The error bar = SEM. Data was statistically analyzed via one-way ANOVA (Dunnet’s multiple comparisons test, *p<0.05, ****p<0.0001). **C-F.** BV2 cells were treated with DMSO or antipsychotics and followed by incubated with 250 nM Aβ40-HyLite-647 and 75 nM LysoTracker DND-99 for an hour, fixed with PFA 4% and imaged on a Nikon A1R. (C) Representative image of Aβ uptake assay. Aβ40 in green and nucleus were stained with Höchst (Blue). Scale bar is 30 μm. (D) Quantification of SFigure 3C. Data was statistically analyzed via one-way ANOVA (Dunnet’s multiple comparisons test, *p<0.05, **p<0.005). n= 50-70 cells per conditions. (E) Representative image of LysoTracker (red). Nuclei were stained with Höchst (Blue). Scale bar is 30 μm. (F) Quantification of SFigure 3E. Data was statistically analyzed via one-way ANOVA (Dunnet’s multiple comparisons test, **p<0.005, ****p<0.0001). n= 50-70 cells per conditions.

**SFigure 4. Antipsychotics alter microglia mediated synaptosome uptake uptake. A-C**. BV2 cells were treated with DMSO or antipsychotics and followed by incubated with ^td^Tomato-tagged synaptosomes for an hour, fixed with PFA 4% and imaged using a Nikon A1R. Cytochalasin-D treatment was used as negative control. (A) Representative image of uptaken synaptosomes (red. Nuclei were stained with Höchst (Blue). Scale bar is 20 μm. (B,C) Quantification of SFigure 4A. Data was statistically analyzed via one-way ANOVA (Dunnet’s multiple comparisons test, ns= not significant, **p<0.005, **p<0.005, ****p<0.0001). n= 50-70 cells per conditions.

