## Supplementary Data for "Olanzapine, Risperidone and Clozapine prescribing is associated with increased risk for Alzheimer’s Disease reflecting antipsychotic-specific effects on microglial phagocytosis"

**Supplementary Methods**

| **Neuronal maintenance medium** | | | |
| --- | --- | --- | --- |
| **Product name** | **Supplier** | **Catalogue number** | **Amount for 50 mL** |
| DMEM:F12 -glutamax | Life Technologies | 11330032 | 25mL |
| Neurobasal Media Plus | Gibco | A35829-01 | 25mL |
| Insulin (10mg/ml) | Sigma | I9278 | 12.5μl |
| 2-mercaptoethanol (50mM) | Life Technologies | 31350 | 50μl |
| N2 | Life Technologies | 17502048 | 250μl |
| B27 | Life Technologies | 17504044 | 500μl |
| Glutamax | Thermofisher | 35050061 | 500μl |
| Laminin | Sigma | L2020 | 50μl |

**Supplementary Data**

| STable 1. Temporal pattern analyses of the associations between use of antipsychotics during different years before the index date and risk of any dementia, a nationwide nested case-control study in Sweden, July 2005-December 2013* | |
| --- | --- |
| Time of antipsychotics use | OR (95%CI) |
| Olanzapine |  |
| 0-1 year | 3.21 (2.98-3.45) |
| 1-2 year | 2.42 (2.22-2.65) |
| 2-3 year | 2.38 (2.14-2.64) |
| 3-4 year | 2.44 (2.16-2.75) |
| 4-5 year | 2.46 (2.13-2.83) |
| 5-9 year | 2.24 (1.92-2.61) |
| Risperidone |  |
| 0-1 year | 4.52 (4.35-4.69) |
| 1-2 year | 3.24 (3.08-3.42) |
| 2-3 year | 2.86 (2.67-3.07) |
| 3-4 year | 2.75 (2.52-3.00) |
| 4-5 year | 2.77 (2.48-3.10) |
| 5-9 year | 2.28 (2.00-2.59) |
| Clozapine |  |
| 0-1 year | 6.11 (4.81-7.75) |
| 1-2 year | 4.98 (3.81-6.50) |
| 2-3 year | 4.61 (3.44-6.17) |
| 3-4 year | 4.52 (3.17-6.45) |
| 4-5 year | 3.97 (2.54-6.18) |
| 5-9 year | 5.95 (3.64-9.75) |
| Abbreviations: CI, confidence interval; OR, odds ratio. | |
| *Analyses were adjusted for age, sex, and area of residence (matching factors). | |

| STable 2. Sensitivity analyses of the association between use of antipsychotics and risk of any dementia, a nationwide nested case-control study in Sweden, July 2008-December 2013* | | | | | | | | |
| --- | --- | --- | --- | --- | --- | --- | --- | --- |
| Use of antipsychotics |  | Sensitivity analysis †, OR (95%CI) | | |  | Sensitivity analysis ⱡ, OR (95%CI) | | |
|  |  | Olanzapine | Risperidone | Clozapine |  | Olanzapine | Risperidone | Clozapine |
| No use |  | 1.00(reference) | 1.00(reference) | 1.00(reference) |  | 1.00(reference) | 1.00(reference) | 1.00(reference) |
| Use |  | 2.38(2.09-2.70) | 2.72(2.47-3.00) | 5.16(3.45-7.69) |  | 2.28(1.96-2.66) | 2.41(2.12-2.75) | 6.16(3.75-10.14) |
| Low cDDD | | 2.38(1.91-2.96) | 2.59(2.21-3.03) | 5.28(2.67-10.45) |  | 2.12(1.61-2.78) | 2.40(1.93-2.99) | 13.05(5.45-31.26) |
| Medium cDDD | | 2.20(1.76-2.73) | 3.24(2.72-3.87) | 6.86(3.44-13.69) |  | 2.27(1.75-2.96) | 2.63(2.08-3.31) | 5.00(1.98-12.60) |
| High cDDD | | 2.58(2.07-3.22) | 2.46(2.08-2.92) | 3.69(1.81-7.55) |  | 2.47(1.89-3.22) | 2.23(1.77-2.82) | 3.22(1.33-7.82) |
| P for trend | | <0.0001 | <0.0001 | <0.0001 |  | <0.0001 | <0.0001 | <0.0001 |
| Abbreviations: cDDD, cumulative defined daily dose; CI, confidence interval; OR, odds ratio. | | | | | | | | |
| Analyses were adjusted for age, sex, and area of residence (matching factors). | | | | | | | | |
| † Analysis included only antipsychotics dispensed more than **four** years before the index date (date of diagnosis for cases and date of selection for controls). | | | | | | | | |
| ⱡ Analysis included only antipsychotics dispensed more than **five** years before the index date (date of diagnosis for cases and date of selection for controls). | | | | | | | | |

| STable 3. Association between use of antipsychotics and risk of any dementia, a nationwide nested case-control study in Sweden, July 2008-December 2013* | | | | | | | | | |
| --- | --- | --- | --- | --- | --- | --- | --- | --- | --- |
|  |  | Olanzapine | |  | Risperidone | |  | Clozapine | |
| Characteristics |  | Cases/Controls | OR (95%CI) |  | Cases/Controls | OR (95%CI) |  | Cases/Controls | OR (95%CI) |
| Male |  |  |  |  |  |  |  |  |  |
| No use |  | 34741/173733 | 1.00(reference) |  | 34542/173551 | 1.00(reference) |  | 34871/174047 | 1.00(reference) |
| Use |  | 177/353 | 2.50(2.09-3.00) |  | 376/535 | 3.55(3.11-4.06) |  | 47/39 | 6.12(3.99-9.39) |
| Low cDDD |  | 60/109 | 2.74(2.00-3.76) |  | 147/196 | 3.79(3.06-4.70) |  | 20/8 | 12.49(5.50-28.36) |
| Medium cDDD |  | 53/118 | 2.25(1.62-3.11) |  | 98/142 | 3.48(2.69-4.50) |  | 14/15 | 4.74(2.29-9.83) |
| High cDDD |  | 64/126 | 2.54(1.88-3.43) |  | 131/197 | 3.37(2.70-4.21) |  | 13/16 | 4.14(1.99-8.61) |
| P for trend |  |  | <0.0001 |  |  | <0.0001 |  |  | <0.0001 |
| Female |  |  |  |  |  |  |  |  |  |
| No use |  | 50468/251792 | 1.00(reference) |  | 50063/251046 | 1.00(reference) |  | 50774/252476 | 1.00(reference) |
| Use |  | 335/721 | 2.32(2.03-2.64) |  | 740/1467 | 2.53(2.31-2.77) |  | 29/37 | 3.91(2.40-6.35) |
| Low cDDD |  | 107/271 | 1.97(1.58-2.47) |  | 322/636 | 2.54(2.22-2.91) |  |  |  |
| Medium cDDD |  | 122/229 | 2.64(2.12-3.29) |  | 196/360 | 2.73(2.29-3.24) |  |  |  |
| High cDDD |  | 106/221 | 2.40(1.90-3.02) |  | 222/471 | 2.36(2.01-2.77) |  |  |  |
| P for trend |  |  | <0.0001 |  |  | <0.0001 |  |  |  |
| Age<70 years |  |  |  |  |  |  |  |  |  |
| No use |  | 6467/32857 | 1.00(reference) |  | 6503/32909 | 1.00(reference) |  | 6571/32970 | 1.00(reference) |
| Use |  | 131/133 | 4.99(3.91-6.36) |  | 95/81 | 5.95(4.42-8.03) |  | 27/20 | 6.96(3.87-12.53) |
| Low cDDD |  | 45/51 | 4.44(2.97-6.63) |  | 30/14 | 10.88(5.76-20.53) |  |  |  |
| Medium cDDD |  | 35/36 | 5.02(3.14-8.03) |  | 13/17 | 3.82(1.86-7.87) |  |  |  |
| High cDDD |  | 51/46 | 5.58(3.74-8.31) |  | 52/50 | 5.31(3.59-7.86) |  |  |  |
| P for trend |  |  | <0.0001 |  |  | <0.0001 |  |  |  |
| Age>=70 years |  |  |  |  |  |  |  |  |  |
| No use |  | 78742/392668 | 1.00(reference) |  | 78102/391688 | 1.00(reference) |  | 79074/393553 | 1.00(reference) |
| Use |  | 381/941 | 2.02(1.79-2.27) |  | 1021/1921 | 2.67(2.47-2.88) |  | 49/56 | 4.37(2.98-6.41) |
| Low cDDD |  | 122/329 | 1.85(1.50-2.27) |  | 439/818 | 2.70(2.40-3.03) |  |  |  |
| Medium cDDD |  | 140/311 | 2.23(1.83-2.72) |  | 281/485 | 2.91(2.51-3.37) |  |  |  |
| High cDDD |  | 119/301 | 1.98(1.60-2.44) |  | 301/618 | 2.45(2.13-2.81) |  |  |  |
| P for trend |  |  | <0.0001 |  |  | <0.0001 |  |  |  |
| Abbreviations: cDDD, cumulative defined daily dose; CI, confidence interval; OR, odds ratio. | | | | | | | | | |
| *Analysis included only antipsychotics dispensed more than three years before the index date. Analyses were adjusted for age, sex, and area of residence (matching factors). Small sample size in the subgroup analysis of Clozapine precludes the analysis by cumulative defined daily doses. | | | | | | | | | |

| STable 4. Association between use of antipsychotics and risk of Alzheimer’s disease, vascular dementia and other dementias, a nationwide nested case-control study in Sweden, July 2008-December 2013* | | | | | | | | | |
| --- | --- | --- | --- | --- | --- | --- | --- | --- | --- |
|  |  | Olanzapine | |  | Risperidone | |  | Clozapine | |
| Subtypes of dementia |  | Cases/Controls | OR (95%CI) |  | Cases/Controls | OR (95%CI) |  | Cases/Controls | OR (95%CI) |
| Alzheimer's disease |  |  |  |  |  |  |  |  |  |
| No use |  | 25164/125200 | 1.00(reference) |  | 25057/125117 | 1.00(reference) |  | 25234/125518 | 1.00(reference) |
| Use |  | 77/342 | 1.12(0.88-1.44) |  | 184/425 | 2.16(1.82-2.57) |  | 7/24 | 1.45(0.62-3.36) |
| Vascular dementia |  |  |  |  |  |  |  |  |  |
| No use |  | 13970/69944 | 1.00(reference) |  | 13916/69820 | 1.00(reference) |  | 14079/70102 | 1.00(reference) |
| Use |  | 111/170 | 3.26(2.56-4.14) |  | 165/294 | 2.80(2.32-3.40) |  | 2/12 | 0.84(0.19-3.74) |
| Other dementias |  |  |  |  |  |  |  |  |  |
| No use |  | 46075/230381 | 1.00(reference) |  | 45632/229660 | 1.00(reference) |  | 46332/230903 | 1.00(reference) |
| Use |  | 324/562 | 2.88(2.51-3.30) |  | 767/1283 | 3.02(2.76-3.30) |  | 67/40 | 8.53(5.74-12.66) |
| Abbreviations: CI, confidence interval; OR, odds ratio. | | | | | | | | | |
| *Analysis included only antipsychotics dispensed more than three years before the index date (date of diagnosis for cases and date of selection for controls). Analyses were adjusted for age, sex, and area of residence (matching factors). | | | | | | | | | |

| STable 5. Association between use of antipsychotics and risk of any dementia, stratified by clinical diagnosis of schizophrenia or bipolar disorders, a nationwide nested case-control study in Sweden, July 2008-December 2013* | | | | | | | | |
| --- | --- | --- | --- | --- | --- | --- | --- | --- |
|  | Olanzapine | |  | Risperidone | |  | Clozapine | |
| Characteristics | Cases/Controls | OR (95%CI) |  | Cases/Controls | OR (95%CI) |  | Cases/Controls | OR (95%CI) |
| Individuals without schizophrenia or bipolar disorders |  |  |  |  |  |  |  |  |
| No | 80315/400409 | 1.00(reference) |  | 79640/399319 | 1.00(reference) |  | 80537/400942 | 1.00(reference) |
| Yes | 273/574 | 2.36 (2.04-2.73) |  | 948/1664 | 2.87 (2.64-3.10) |  | 51/41 | 6.20 (4.11-9.35) |
| Low cDDD | 89/197 | 2.25 (1.75-2.89) |  | 352/588 | 3.02 (2.64-3.44) |  | 24/17 | 7.00 (3.76-13.03) |
| Medium cDDD | 94/188 | 2.48 (1.93-3.17) |  | 309/523 | 2.97 (2.58-3.41) |  | 12/12 | 5.00 (2.25-11.13) |
| High cDDD | 90/189 | 2.36 (1.84-3.04) |  | 287/553 | 2.61 (2.26-3.01) |  | 15/12 | 6.24 (2.92-13.34) |
| P for trend |  | <0.0001 |  |  | <0.0001 |  |  | <0.0001 |
| Individuals with schizophrenia or bipolar disorders |  |  |  |  |  |  |  |  |
| No | 4894/25116 | 1.00(reference) |  | 4965/25278 | 1.00(reference) |  | 5108/25581 | 1.00(reference) |
| Yes | 239/500 | 2.40 (2.05-2.80) |  | 168/338 | 2.50 (2.07-3.01) |  | 25/35 | 3.62 (2.16-6.07) |
| Low cDDD | 74/173 | 2.14 (1.63-2.81) |  | 64/125 | 2.57 (1.90-3.47) |  | 10/10 | 5.11 (2.13-12.30) |
| Medium cDDD | 82/168 | 2.45 (1.88-3.19) |  | 52/99 | 2.64 (1.88-3.69) |  | 9/12 | 3.85 (1.62-9.16) |
| High cDDD | 83/159 | 2.62 (2.01-3.42) |  | 52/114 | 2.30 (1.65-3.19) |  | 6/13 | 2.31 (0.88-6.07) |
| P for trend |  | <0.0001 |  |  | <0.0001 |  |  | <0.0001 |
| Abbreviations: cDDD, cumulative defined daily dose; CI, confidence interval; OR, odds ratio. | | | | | | | | |
| * Analysis included only antipsychotics dispensed more than three years before the index date (date of diagnosis for cases and date of selection for controls). Analyses were adjusted for age, sex, and area of residence (matching factors). | | | | | | | | |

| STable 6. Diagnoses of psychotic disorders among individuals with antipsychotic treatment but without a diagnosis of schizophrenia or bipolar disorders, stratified by having dementia or not, a nationwide nested case-control study in Sweden, July 2005-December 2013* | | | | | | | | | |
| --- | --- | --- | --- | --- | --- | --- | --- | --- | --- |
|  |  | Olanzapine | |  | Risperidone | |  | Clozapine | |
| Psychotic disorders |  | Individuals with dementia, % (n=2417) | Controls, % (n=3276) |  | Individuals with dementia, % (n=15306) | Controls, % (18785) |  | Individuals with dementia, % (n=334) | Controls, % (n=214) |
| Anxiety disorders |  | 369 (15.27) | 633 (19.32) |  | 788 (5.15) | 1053 (5.61) |  | 33 (9.88) | 18 (8.41) |
| Autism spectrum disorders |  | 2 (0.08) | 5 (0.15) |  | 5 (0.03) | 6 (0.03) |  | 0 (0.00) | 0 (0.00) |
| Conduct disorders |  | 20 (0.83) | 7 (0.21) |  | 57 (0.37) | 38 (0.20) |  | 4 (1.20) | 2 (0.93) |
| Depressive disorders |  | 639 (26.44) | 1038 (31.68) |  | 1577 (10.30) | 1801 (9.59) |  | 49 (14.67) | 21 (9.81) |
| Eating disorders |  | 5 (0.21) | 9 (0.27) |  | 7 (0.05) | 8 (0.04) |  | 0 (0.00) | 0 (0.00) |
| Personality disability |  | 37 (1.53) | 55 (1.68) |  | 72 (0.47) | 71 (0.38) |  | 1 (0.30) | 1 (0.47) |
| Substance use disorders |  | 149 (6.16) | 130 (3.97) |  | 347 (2.27) | 261 (1.39) |  | 8 (2.40) | 1 (0.47) |
| * Psychotic disorders were identified from three years before the first date of antipsychotics dispense to three years after the last date of antipsychotics dispense. | | | | | | | | | |

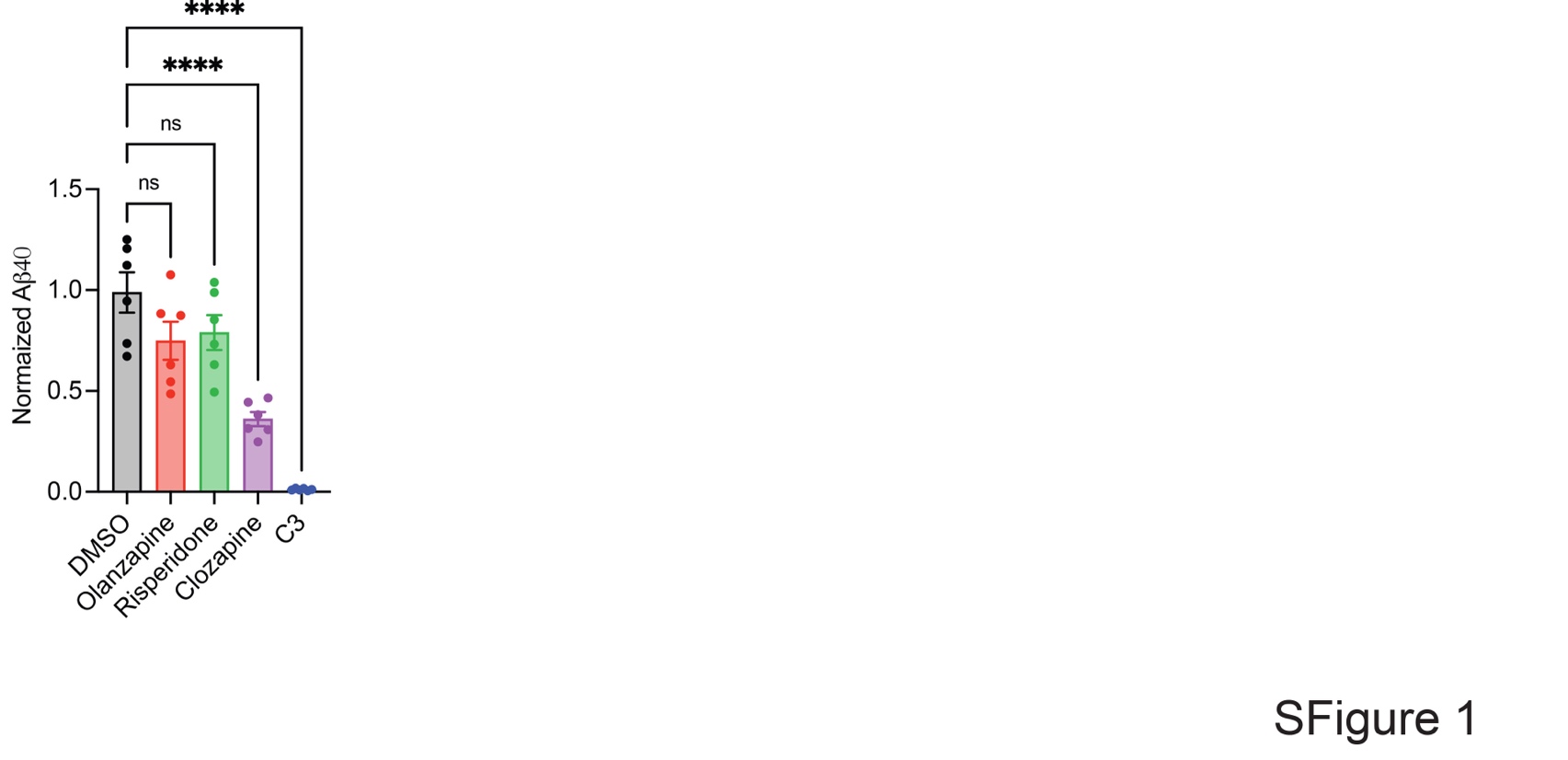

**SFigure 1. Clozapine reduces secreted Aβ levels in HeLa swAPP cells.** HeLa swAPP cells were treated with Olanzapine (10μM), Risperidone (1 μM) and Clozapine (25 μM) for 24 hours and the medium were collected for 4h to analyze the quantity of Aβ using electrochemiluminescence assay (MSD) and the count normalized to total protein level in cell lysate. DMSO was used as control. The error bar = SEM. Data was statistically analyzed via one-way ANOVA (Dunnet’s multiple comparisons test, ns= not significant, ****p<0.0001).

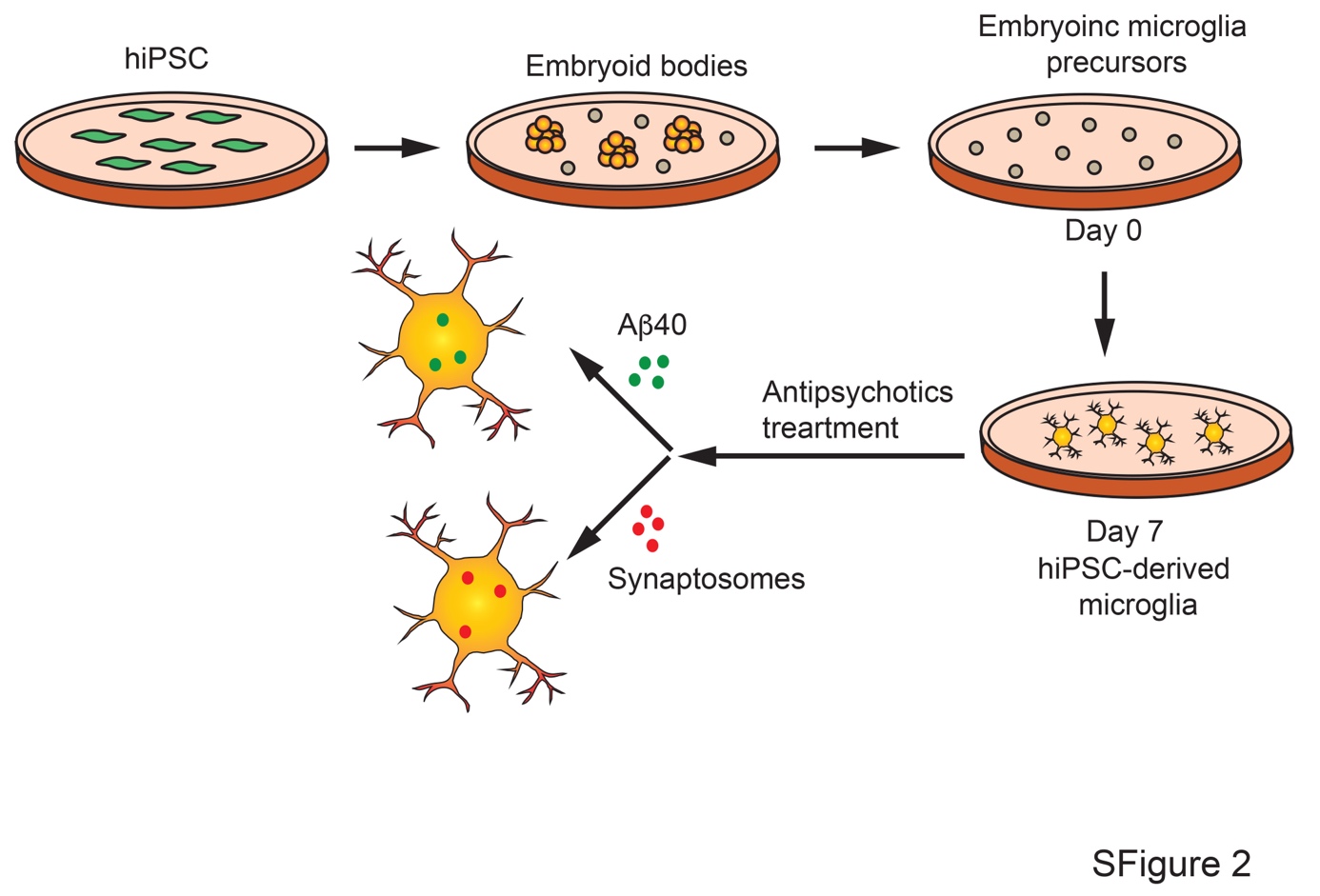

**SFigure 2.** Schematic diagram of hiPSC derived microglia preparation and experimental plan.

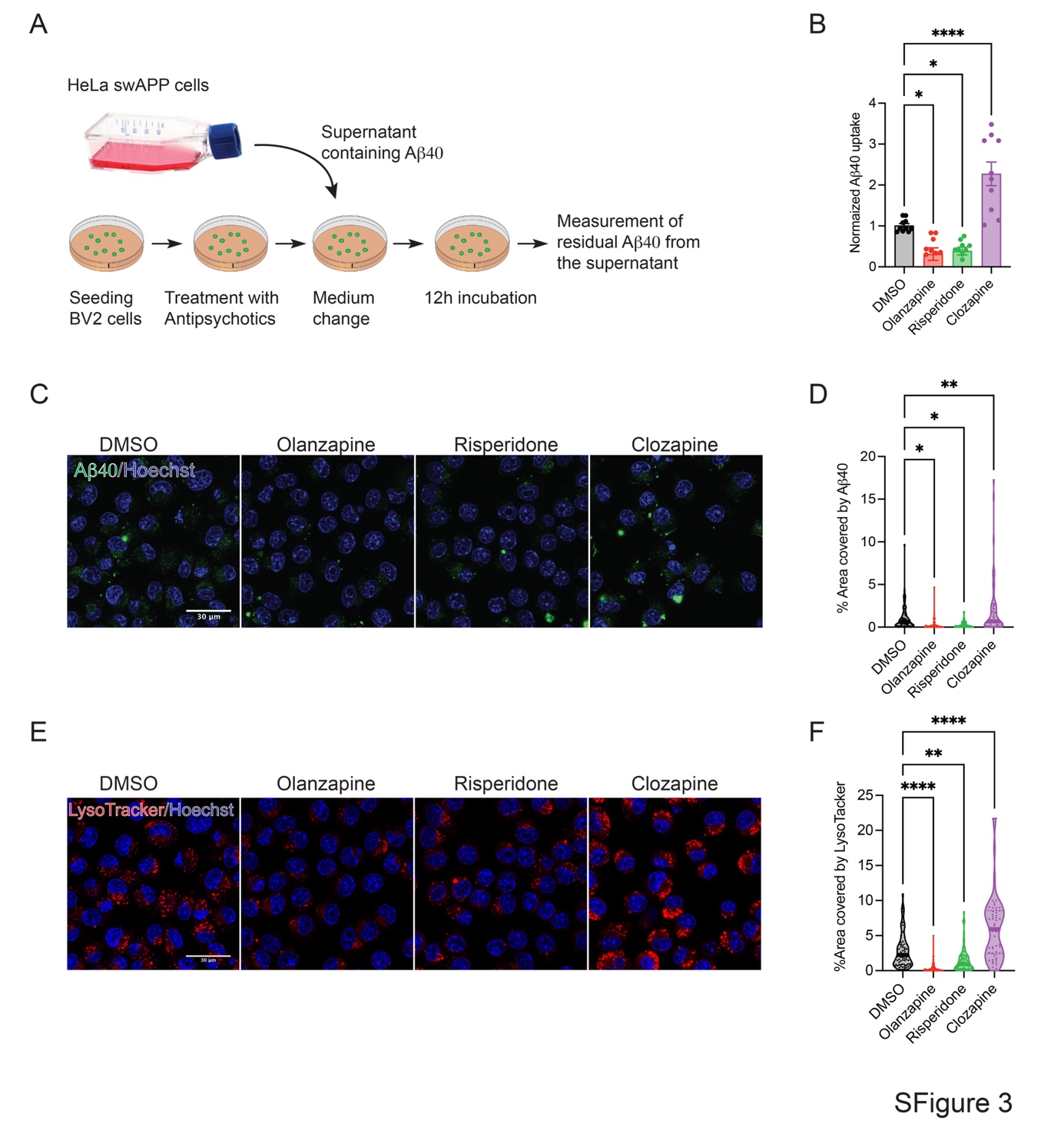

**SFigure 3. Antipsychotics alter microglia mediated Aβ40 uptake. A.** Experimental overview of Aβ uptake assay. **B.** BV2 microglia cells were treated with DMSO or antipsychotics (Olanzapine: 10 μM, Risperidone: 1 μM and Clozapine: 25 μM) for 10h followed by incubated overnight with supernatant taken from HeLa (swAPP) containing Aβ. The residual Aβ was quantified using MSD and normalized to total protein. Uptake Aβ was quantified through calculation the change from initial, control loaded Aβ amounts. The error bar = SEM. Data was statistically analyzed via one-way ANOVA (Dunnet’s multiple comparisons test, *p<0.05, ****p<0.0001). **C-F.** BV2 cells were treated with DMSO or antipsychotics and followed by incubated with 250 nM Aβ40-HyLite-647 and 75 nM LysoTracker DND-99 for an hour, fixed with PFA 4% and imaged on a Nikon A1R. (C) Representative image of Aβ uptake assay. Aβ40 in green and nucleus were stained with Höchst (Blue). Scale bar is 30 μm. (D) Quantification of SFigure 3C. Data was statistically analyzed via one-way ANOVA (Dunnet’s multiple comparisons test, *p<0.05, **p<0.005). n= 50-70 cells per conditions. (E) Representative image of LysoTracker (red). Nuclei were stained with Höchst (Blue). Scale bar is 30 μm. (F) Quantification of SFigure 3E. Data was statistically analyzed via one-way ANOVA (Dunnet’s multiple comparisons test, **p<0.005, ****p<0.0001). n= 50-70 cells per conditions.

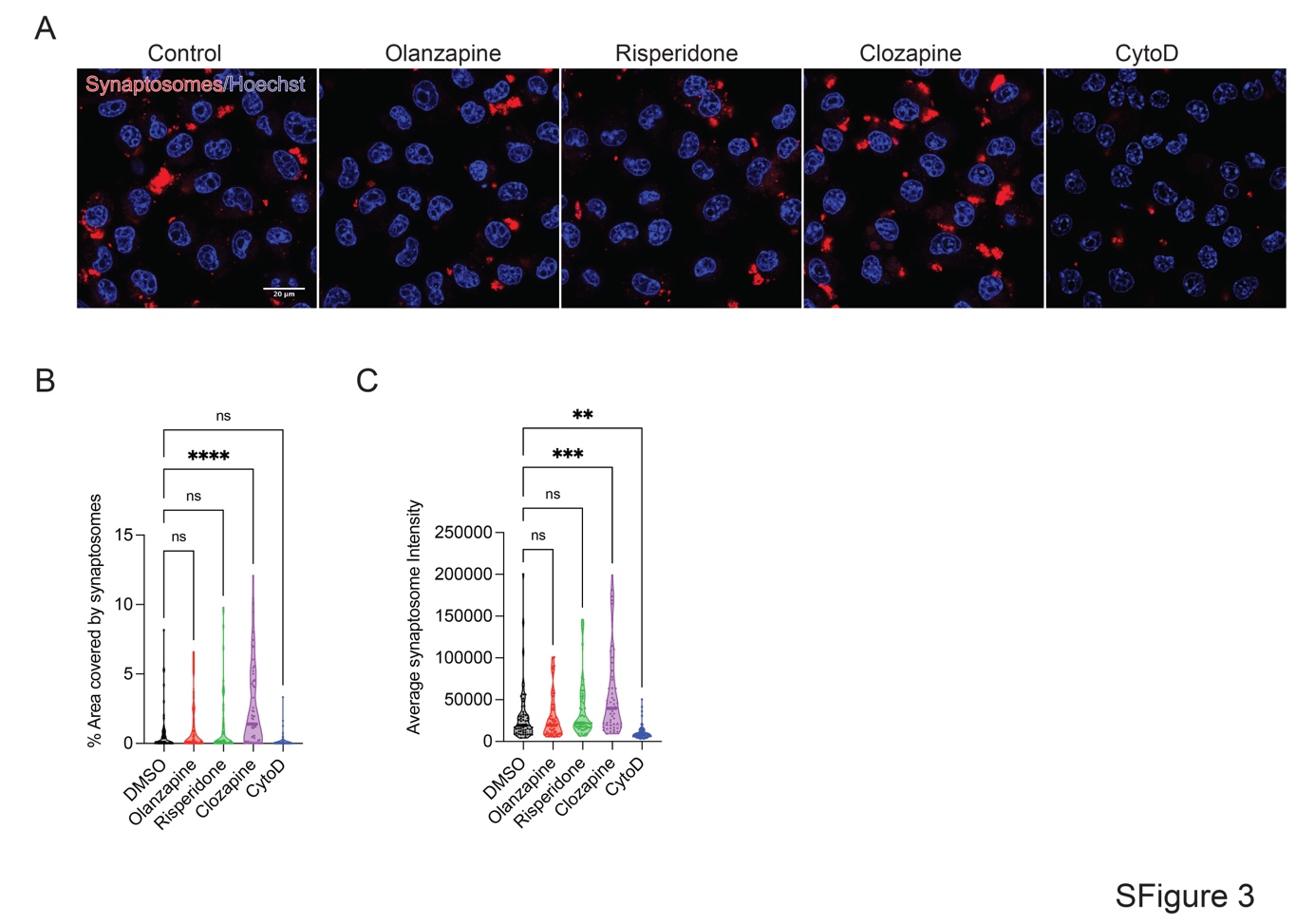

**SFigure 4. Antipsychotics alter microglia mediated synaptosome uptake uptake. A-C**. BV2 cells were treated with DMSO or antipsychotics and followed by incubated with ^td^Tomato-tagged synaptosomes for an hour, fixed with PFA 4% and imaged using a Nikon A1R. Cytochalasin-D treatment was used as negative control. (A) Representative image of uptaken synaptosomes (red. Nuclei were stained with Höchst (Blue). Scale bar is 20 μm. (B,C) Quantification of SFigure 4A. Data was statistically analyzed via one-way ANOVA (Dunnet’s multiple comparisons test, ns= not significant, **p<0.005, **p<0.005, ****p<0.0001). n= 50-70 cells per conditions.
